# “Characteristics and Predictors of Pediatric and Adult Patients with Inherited Retinal Degenerations at Time of Presentation: Tertiary Care Ophthalmology Clinic Data”

**DOI:** 10.1101/2024.08.15.24312074

**Authors:** Matthew T. McLaughlin, Caleb P. Ganansky, Ayman W. Taher, Melissa A. Trudrung, William Van De Car, Jonathan Le, Kyle D. Peterson, Kimberly E. Stepien, Melanie A. Schmitt

## Abstract

Inherited Retinal Degenerations (IRDs) are a group of diseases where genetic variants lead to retinal photoreceptor dysfunction and subsequent visual impairment. We aimed to compare the characteristics of pediatric versus adult IRD patients at the time of presentation at a tertiary care IRD clinic. A retrospective chart review of 538 patients with IRDs was conducted. Information obtained included age, diagnosis, presenting characteristics, demographics, distance from the clinic, and referring physician. This study found that high hyperopia, congenital syndactyly, high refractive error, high astigmatism, and a history of developmental delay were most predictive of pediatric presentation. In adults, we found reduced central vision, peripheral vision loss, color vision deficits, nyctalopia, flashes/floaters, cataracts, diabetes mellitus, obesity, cardiac conditions, and a family history of cataract were most predictive of presentation. There was a greater proportion of pediatric patients presenting from 20 or more miles away. Additionally, there was no significant difference between the type of healthcare provider referring adult or pediatric patients. This study identifies characteristics predictive of pediatric and adult presentation in IRD patients thus addressing current knowledge gaps. A better understanding of these characteristics may provide for quicker recognition, education of clinicians likely to encounter IRDs, and allow for earlier treatment.

## Introduction

Inherited Retinal Degenerations (IRDs) represent a group of diseases where genetic variants lead to retinal photoreceptor dysfunction and subsequent visual impairment and blindness. IRDs can present in the first year of life to adulthood and vary in the rapidity and severity of progression. To date, over 280 genes have been implicated in IRD pathogenesis each causing different clinical phenotypes^1^. Given the diversity in presentation of IRDs, early identification and diagnosis in patients proves challenging.

IRDs are commonly associated with refractive errors, particularly myopia^2^. Furthermore, early-onset high myopia has been consistently linked with IRDs including retinitis pigmentosa (RP) caused by *RPGR* and *RP1* mutations^3,4,5,6^, congenital stationary night blindness^7^, and cone dystrophy and cone-rod dystrophy^8^. High hyperopia has also been linked to IRDs including RP^4,9^ and other dystropies^2^. Though the association between IRDs and refractive errors like high myopia are well established, the relationship between IRDs and high hyperopia is not as well described in the literature.

Gene-based therapies represent an exciting area of research for treatment of IRDs with known monogenic variants. In 2017, voretigene neparvovec-rzyl became the first gene therapy approved by the US Food and Drug Administration to treat an IRD in 2017^10^. This landmark achievement spurred significant international investment in research for other IRD gene therapeutics. Despite these exciting advancements, an analysis of the current knowledge of IRDs recognized that the natural history and environmental factors are areas in need of continued research^11^. Significant knowledge gaps remain in our understanding of patient factors that contribute to early or delayed presentation for treatment at tertiary care centers.

The primary objective of this study was to investigate the characteristics of pediatric versus adult IRD patients at the time of presentation at the IRD Clinic at the University of Wisconsin Department of Ophthalmology and Visual Sciences. We hypothesized that poor visual acuity and family history would be the most predictive of early recognition of IRDs and that living closer to a tertiary care center would be associated with a younger patient age of referral compared to patients living further away from the tertiary care center.

## Materials and Methods

### Patient Inclusion

After Institutional Review Board approval under protocol number 2020-1207, we performed a retrospective cohort study including patients of all ages who were diagnosed with an IRD at a single tertiary ophthalmology referral center (the Inherited Retinal Degeneration Clinic at the University of Wisconsin Department of Ophthalmology and Visual Sciences) between January 1, 1990 and January 1, 2020. All patients who presented to the clinic with IRDs who agreed to participate were added to an IRB approved REDCap database. Inclusion criteria included anyone referred to the tertiary care center with a diagnosis of IRD who agreed to be placed in the REDCap database. Exclusion criteria included anyone whose initial visit at the clinic was not with one of the study’s IRD ophthalmologists, anyone who did not agree to be placed into the REDCap database, and those who were not diagnosed with IRDs.

### Data Collection

A manual chart review was performed to collect demographic information, relevant personal and family medical history, IRD diagnosis, disease, and treatment information from a single patient encounter using a standardized interview template. Demographic information included age, sex, race (White, Black, Asian, Hispanic, Native American, Other), education (High School or less, Bachelors, Graduate, N/A), driving status, employment status, and home address. Each patient’s home address and the address of the eye clinic where the visit was conducted were used to estimate commute distance (<20, 20-80, >80 miles) with Google Maps (Google LLC, Mountain View, Calif.). IRD information included ocular diagnosis, syndrome, mutation gene name(s), left and right eye distance visual acuity, left and right eye logMAR scores, and left and right eye lens statuses (phakic – no cataract, phakic – cataract, psuedophakic), referring provider (ophthalmologist, optometrist, primary care physician, emergency department physician, or other), and if the patient previously saw a low vision specialist. IRD signs and symptoms were recorded for each patient including high hyperopia, high myopia, high astigmatism, wears correction, central vision loss, peripheral vision loss, nyctalopia, photophobia, color vision deficits, flashes or floaters, ptosis, cataracts, nystagmus, retinal abnormalities, functional vision loss, strabismus, or amblyopia. Additional past medical information was recorded including presence of obesity, diabetes, cardiac problems, hearing loss, developmental delay, intellectual disability, neurological conditions, abnormal teeth, or if the patient was born prematurely, had congenital renal malformations, polydactyly, or syndactyly. Patient family history information recorded included diagnosis of a family member with an IRD, nystagmus, high refractive error, nyctalopia, vision loss, color vision deficit, polydactyly, syndactyly, cataracts, strabismus, amblyopia, cardiac conditions, or neurological conditions. If any of the aforementioned signs, symptoms, family, or personal history were not present in the electronic health record, they were recorded as negative.

### Outcomes

The primary outcome was to compare the characteristics of pediatric versus adult IRD patients at the time of presentation at the Inherited Retinal Degeneration Clinic at the Department of Ophthalmology and Visual Sciences. The secondary outcome was to investigate differences between pediatric and adult patients with IRD in proximity to the tertiary care center, referring provider, and diagnoses.

### Statistical Analysis

We employed Fisher’s exact tests to analyze all categorical covariates to determine the presence of significant associations between pediatric and adult clinical characteristics. When significance was detected in tables larger than 2×2, we applied the Hommel post-hoc adjustment to correct for multiple comparisons. Additionally, we quantified significant associations by calculating odds ratios and their corresponding 95% confidence intervals to provide a more detailed understanding of the relationship within our data.

## Results

### Patient Characteristics

538 patients with IRDs were identified. Of the 538 patients, 11 were excluded. Data was collected on 527 patients of which 124 were pediatric and 403 were adult. Patient characteristics are further detailed in Table 1.

**Table 1.** Patient characteristics. All NA, other, declined, were removed for analyses.

|  |  | Pediatric (< 18 yo) | Adult (≥ 18 yo) | Total | *p*-value |
| --- | --- | --- | --- | --- | --- |
| *N* |  | 124 | 403 | 527 |  |
| Age |  | 9 (5) | 49 (17) | 39 (22) |  |
| Sex |  |  |  |  |  |
|  | Male | 79 | 212 | 291 | 0.039 |
|  | Female | 45 | 190 | 235 |  |
|  | NA | 1 | 1 | 1 |  |
| Race/Ethnicity |  |  |  |  |  |
|  | White | 86 | 325 | 411 | 0.30 |
|  | Black/African American | 10 | 18 | 28 |  |
|  | Asian | 3 | 6 | 9 |  |
|  | AIAN | 3 | 2 | 5 |  |
|  | Other | 5 | 8 | 10 |  |
|  | Declined | 0 | 1 | 1 |  |
|  | NA | 13 | 43 | 63 |  |
| Patient Drives |  |  |  |  |  |
|  | Yes | 4 | 180 | 184 | < 0.001 |
|  | No | 76 | 190 | 266 |  |
|  | NA | 1 | 13 | 14 |  |
| Referring Provider |  |  |  |  |  |
|  | Ophthalmologist | 51 | 181 | 232 | 0.36 |
|  | Optometrist | 13 | 71 | 84 |  |
|  | Primary Care | 22 | 72 | 94 |  |
|  | Emergency | 0 | 0 | 0 |  |
|  | Other | 38 | 74 | 112 |  |
|  | Unknown | 0 | 5 | 5 |  |

### Characteristics Predictive of Pediatric Presentation

Of the 124 pediatric patients, the ocular characteristics at presentation included high hyperopia (p < 0.001), high myopia (p = 0.004), high astigmatism (p < 0.001), ptosis (p = 0.022), nystagmus (p < 0.001), and strabismus (p = 0.001) (Table 2). The medical characteristics predictive of pediatric presentation were syndactyly (p = 0.041), history of developmental delay (p < 0.001), and history of intellectual disability (p < 0.001) (Table 3). Having a family history of nystagmus (p < 0.001), high refractive error (p = 0.002), vision loss (p < 0.001), strabismus (p = 0.002), and amblyopia (p = 0.045) was predictive of pediatric presentation (Table 4).

**Table 2.** Pediatric and adult patient ocular characteristics at presentation (where P and N denote positive and negative, respectively).

| | Pediatric (< 18 yo) | | Adult (≥ 18 yo) | | Total | | *p*-value |
| --- | --- | --- | --- | --- | --- | --- | --- |
|  | P | N | P | N | P | N |  |
| High Hyperopia | 16 (13%) | 107 (87%) | 1 (0%) | 377 (100%) | 17 (3%) | 484 (97%) | < 0.001 |
| High Myopia | 29 (24%) | 93 (76%) | 47 (12%) | 331 (88%) | 76 (15%) | 424 (85%) | 0.004 |
| High Astigmatism | 29 (24%) | 93 (76%) | 18 (5%) | 360 (95%) | 47 (9%) | 453 (91%) | < 0.001 |
| Wears Correction | 81 (70%) | 40 (30%) | 299 (75%) | 98 (25%) | 380 (73%) | 138 (27%) | 0.078 |
| Reduced Central Vision | 36 (33%) | 72 (67%) | 196 (50%) | 196 (50%) | 232 (46%) | 268 (54%) | 0.002 |
| Peripheral Vision Loss | 27 (25%) | 82 (75%) | 158 (40%) | 235 (60%) | 185 (37%) | 317 (63%) | 0.003 |
| Color Vision Deficits | 37 (34%) | 73 (66%) | 197 (50%) | 196 (50%) | 234 (47%) | 269 (53%) | 0.002 |
| Functional Vision Loss | 0 (0%) | 119 (100%) | 0 (0%) | 397 (100%) | 0 (0%) | 516 (100%) | 1 |
| Nyctalopia | 41 (37%) | 71 (63%) | 256 (65%) | 139 (35%) | 297 (59%) | 210 (41%) | < 0.001 |
| Photophobia | 63 (53%) | 56 (47%) | 240 (61%) | 155 (39%) | 303 (59%) | 211 (41%) | 0.138 |
| Flashes/Floaters | 21 (19%) | 87 (81%) | 257 (66%) | 135 (34%) | 278 (56%) | 222 (44%) | < 0.001 |
| Cataracts | 7 (6%) | 113 (94%) | 212 (53%) | 188 (47%) | 219 (42%) | 301 (58%) | < 0.001 |
| Ptosis | 6 (5%) | 110 (95%) | 5 (1%) | 384 (99%) | 11 (2%) | 494 (98%) | 0.022 |
| Nystagmus | 47 (39%) | 74 (61%) | 43 (11%) | 351 (91%) | 90 (17%) | 425 (83%) | < 0.001 |
| Retinal Abnormalities | 117 (98%) | 3 (2%) | 394 (99%) | 3 (1%) | 511 (99%) | 6 (1%) | 0.141 |
| Both eyes affected | 120 (99%) | 1 (1%) | 391 (99%) | 3 (1%) | 511 (99%) | 4 (1%) | 1 |
| Strabismus | 16 (13%) | 107 (87%) | 17 (4%) | 374 (96%) | 33 (6%) | 481 (94%) | 0.001 |
| Amblyopia | 11 (9%) | 113 (91%) | 18 (5%) | 374 (95%) | 29 (5%) | 487 (95%) | 0.077 |

**Table 3.** Pediatric and adult patient additional medical characteristics at presentation (where P and N denote positive and negative, respectively).

|  | Pediatric (< 18 yo) |  | Adult (≥ 18 yo) |  | Total |  | *p*-value |
| --- | --- | --- | --- | --- | --- | --- | --- |
|  | P | N | P | N | P | N |  |
| Congenital |  |  |  |  |  |  |  |
| Polydactyly | 0 (0%) | 115 (100%) | 12 (3%) | 382 (97%) | 12 (2%) | 497 (98%) | 0.078 |
| Syndactyly | 3 (3%) | 112 (97%) | 1 (0%) | 393 (100%) | 4 (1%) | 504 (99%) | 0.041 |
| Kidney Malfunction | 0 (0%) | 118 (100%) | 11 (3%) | 380 (97%) | 11 (2%) | 498 (98%) | 0.076 |
| History of Prematurity | 14 (12%) | 102 (88%) | 30 (8%) | 360 (92%) | 44 (9%) | 462 (91%) | 0.187 |
| History of<br>Developmental Delay | 15 (13%) | 103 (87%) | 9 (2%) | 382 (98%) | 24 (5%) | 485 (95%) | < 0.001 |
| History of Intellectual<br>Disability | 16 (14%) | 102 (86%) | 17 (4%) | 375 (96%) | 33 (6%) | 477 (94%) | < 0.001 |
| Abnormal Teeth | 2 (0%) | 116 (100%) | 1 (0%) | 393 (100%) | 3 (1%) | 509 (99%) | 0.134 |
| Diabetes Mellitus | 1 (0%) | 117 (100%) | 41 (10%) | 356 (90%) | 42 (8%) | 473 (92%) | < 0.001 |
| Obesity | 9 (8%) | 109 (92%) | 77 (20%) | 316 (80%) | 86 (17%) | 425 (83%) | 0.002 |
| Cardiac Condition | 7 (6%) | 117 (94%) | 100 (26%) | 288 (74%) | 107 (20%) | 405 (80%) | < 0.001 |
| Neurologic Condition | 19 (16%) | 99 (84%) | 75 (19%) | 319 (81%) | 94 (18%) | 418 (82%) | 0.502 |
| Hearing Loss | 19 (16%) | 99 (84%) | 89 (23%) | 301 (77%) | 108 (21%) | 400 (79%) | 0.125 |

**Table 4.** Pediatric and adult patient family history at presentation (where P and N denote positive and negative, respectively).

| | Pediatric (< 18 yo) | | Adult (≥ 18 yo) | | Total | | *p*-value |
| --- | --- | --- | --- | --- | --- | --- | --- |
|  | P | N | P | N | P | N |  |
| Diagnosed IRD | 33 (27%) | 88 (73%) | 137 (35%) | 257 (65%) | 170 (33%) | 345 (67%) | 0.151 |
| Nystagmus | 13 (11%) | 107 (89%) | 10 (3%) | 378 (97%) | 23 (5%) | 485 (95%) | < 0.001 |
| High Refractive Error | 8 (7%) | 113 (93%) | 4 (1%) | 382 (99%) | 12 (2%) | 495 (98%) | 0.002 |
| Nyctalopia | 16 (13%) | 105 (87%) | 35 (9%) | 352 (91%) | 51 (10%) | 457 (90%) | 0.224 |
| Vision Loss | 50 (75%) | 17 (25%) | 167 (43%) | 224 (57%) | 217 (42%) | 295 (58%) | < 0.001 |
| Color Vision Deficit | 11 (9%) | 110 (91%) | 27 (7%) | 361 (93%) | 38 (7%) | 471 (93%) | 0.432 |
| Polydactyly/Syndactyly | 1 (0%) | 120 (100%) | 6 (2%) | 383 (98%) | 7 (1%) | 503 (99%) | 1 |
| Cataract | 20 (16%) | 103 (84%) | 105 (27%) | 287 (73%) | 125 (24%) | 390 (76%) | 0.022 |
| Strabismus | 28 (24%) | 91 (76%) | 46 (12%) | 349 (88%) | 74 (14%) | 440 (86%) | 0.002 |
| Amblyopia | 17 (15%) | 100 (85%) | 31 (8%) | 364 (82%) | 48 (9%) | 464 (91%) | 0.045 |
| Cardiac Condition | 9 (8%) | 109 (92%) | 46 (12%) | 345 (88%) | 55 (11%) | 454 (89%) | 0.239 |
| Neurological Condition | 15 (12%) | 108 (88%) | 63 (16%) | 329 (84%) | 77 (15%) | 437 (85%) | 0.317 |

Of these variables, the strongest strength of association was found with high hyperopia (Odds Ratio (OR) 49.34, 95% CI 9.89-1195.80), congenital syndactyly (OR 9.62, 95% CI 1.10-227.57), high refractive error (OR 6.60, 95% CI 2.00-26.03), high astigmatism (OR 6.19, 95% CI 3.31-11.86), and a history of developmental delay (OR 6.11, 95% CI 2.62-15.08) (Table 5).

**Table 5.** Univariate odds ratios to quantify associations for each significant covariate.

| Covariate | Odds Ratio | 95% CI |
| --- | --- | --- |
| High Hyperopia | 49.34 | [9.89, 1195.80] |
| High Myopia | 2.20 | [1.30, 3.67] |
| High Astigmatism | 6.19 | [3.31, 11.86] |
| Reduced Central Vision | 0.50 | [0.32, 0.78] |
| Peripheral Vision Loss | 0.49 | [0.30, 0.79] |
| Color Vision Deficits | 0.51 | [0.32, 0.78] |
| Nyctalopia | 0.31 | [0.20, 0.49] |
| Flashes/Floaters | 0.13 | [0.07, 0.21] |
| Cataracts | 0.06 | [0.02, 0.12] |
| Ptosis | 4.16 | [1.20, 15.12] |
| Nystagmus – at presentation | 5.15 | [3.19, 8.41] |
| Strabismus | 3.28 | [1.58, 6.78] |
| Congenital Syndactyly | 9.62 | [1.10, 227.57] |
| History of Developmental Delay | 6.11 | [2.62, 15.08] |
| History of Intellectual Disability | 3.45 | [1.66, 7.14] |
| Diabetes Mellitus | 0.08 | [0.004, 0.39] |
| Obesity | 0.34 | [0.15, 0.68] |
| Cardiac Condition | 0.18 | [0.07, 0.37] |
| Nystagmus – family history | 4.56 | [1.93, 11.07] |
| High Refractive Error | 6.60 | [2.00, 26.03] |
| Vision Loss | 3.91 | [2.21, 7.23] |
| Cataract – family history | 0.53 | [0.31, 0.89] |
| Strabismus – family history | 2.33 | [1.37, 3.93] |
| Amblyopia | 2.00 | [1.04, 3.73] |

### Characteristics Predictive of Adult Presentation

Of the 403 adults, the ocular characteristics most predictive of adult presentation were reduced central vision (p = 0.002), peripheral vision loss (p = 0.003), color vision deficits (p = 0.002), nyctalopia (p < 0.001), flashes/floaters (p < 0.001), and cataracts (p < 0.001) (Table 2). The medical characteristics predictive of adult presentation were diabetes mellitus (p < 0.001), obesity (p = 0.002), and a cardiac condition (p < 0.001) (Table 3). Additionally, A family history of cataract (p = 0.022) was predictive of adult presentation (Table 4).

### Proximity to Tertiary Care Center

There was a smaller proportion of pediatric patients that lived <20 miles (95% CI 8.7-19.7) from the ophthalmology tertiary care center compared to patients that lived 20-80 miles (95% CI 22.0-36.1) away and >80 miles (95% CI 21.9-35.0) away (Figure 1). There was no difference in proportion of pediatric patients between patients living between 20-80 miles (95% CI 22.0-36.1) away as compared to >80 miles (95% CI 21.9-35.0) away from the ophthalmology tertiary care center (Figure 1).

**Figure 1.**
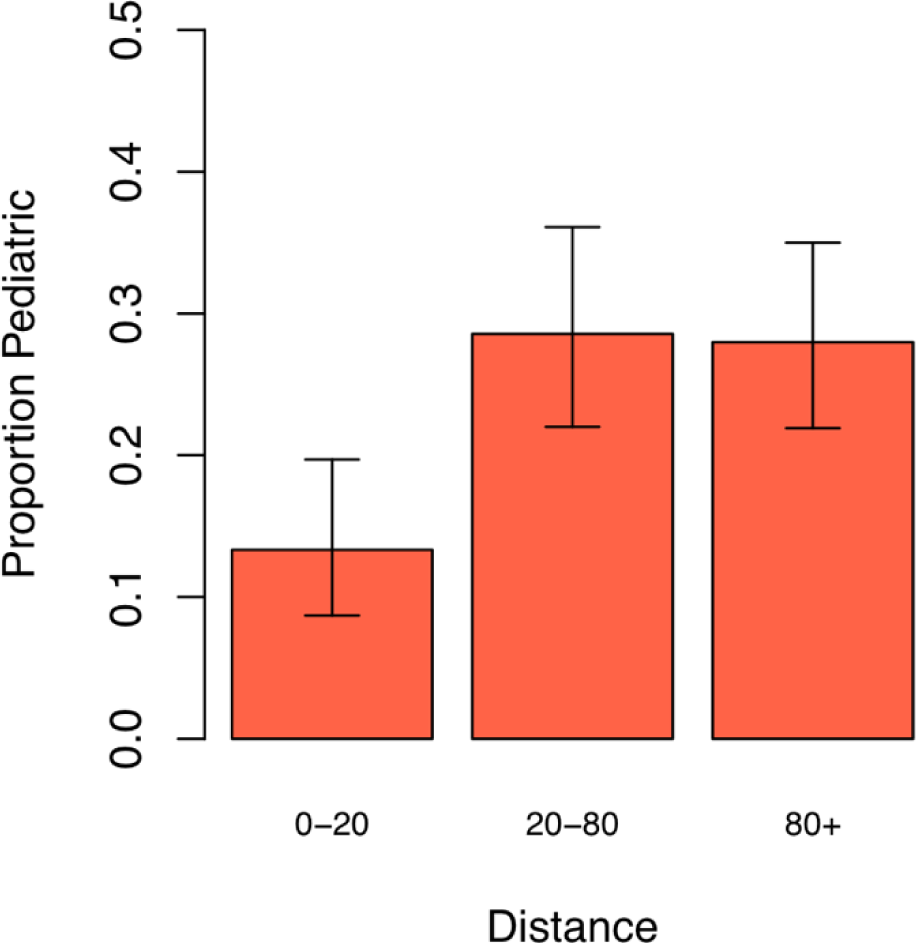
Proportion of pediatric IRD patients in relation to proximity of home to tertiary ophthalmology care center where the error bars denote 95% confidence intervals for the proportions. A distance of 0-20 signifies values less than 20 while 20-80 denotes values 20 or more miles away.

### Referring Provider

There was no significant difference between the referring provider and whether the patient was pediatric or adult (p = 0.36) (Table 1). In terms of the absolute number of referrals, the number of ophthalmologists initiating referrals was overall the highest value in both pediatric and adult patients (Table 1). Primary care physicians additionally comprised a number of referrals (Table 1). There was no significant difference in the proportion of pediatric patients with IRDs referred by any type of clinician (Table 6).

**Table 6.** Post-hoc analysis of proportion of pediatric referring provider.

| Referring Provider |  | p-value |
| --- | --- | --- |
|  | Ophthalmologist vs. Optometrist | 0.534 |
|  | Ophthalmologist vs. Primary Care Physician | 0.771 |
|  | Optometrist vs. Primary Care Physician | 0.534 |

### Diagnosis

The highest proportion of IRDs in the pediatric patients were Leber Congenital Amaurosis (LCA) and cone-rod dystrophy (Figure 2 and Table 7). Other ocular diagnoses of choroideremia, RP, Stargardt disease, and vitelliform dystrophy were present at a lower proportion in pediatric patients (Figure 2 and Table 7). The most common IRD diagnoses in the adult group were RP followed by Stargardt disease (Figure 7).

**Figure 2.**
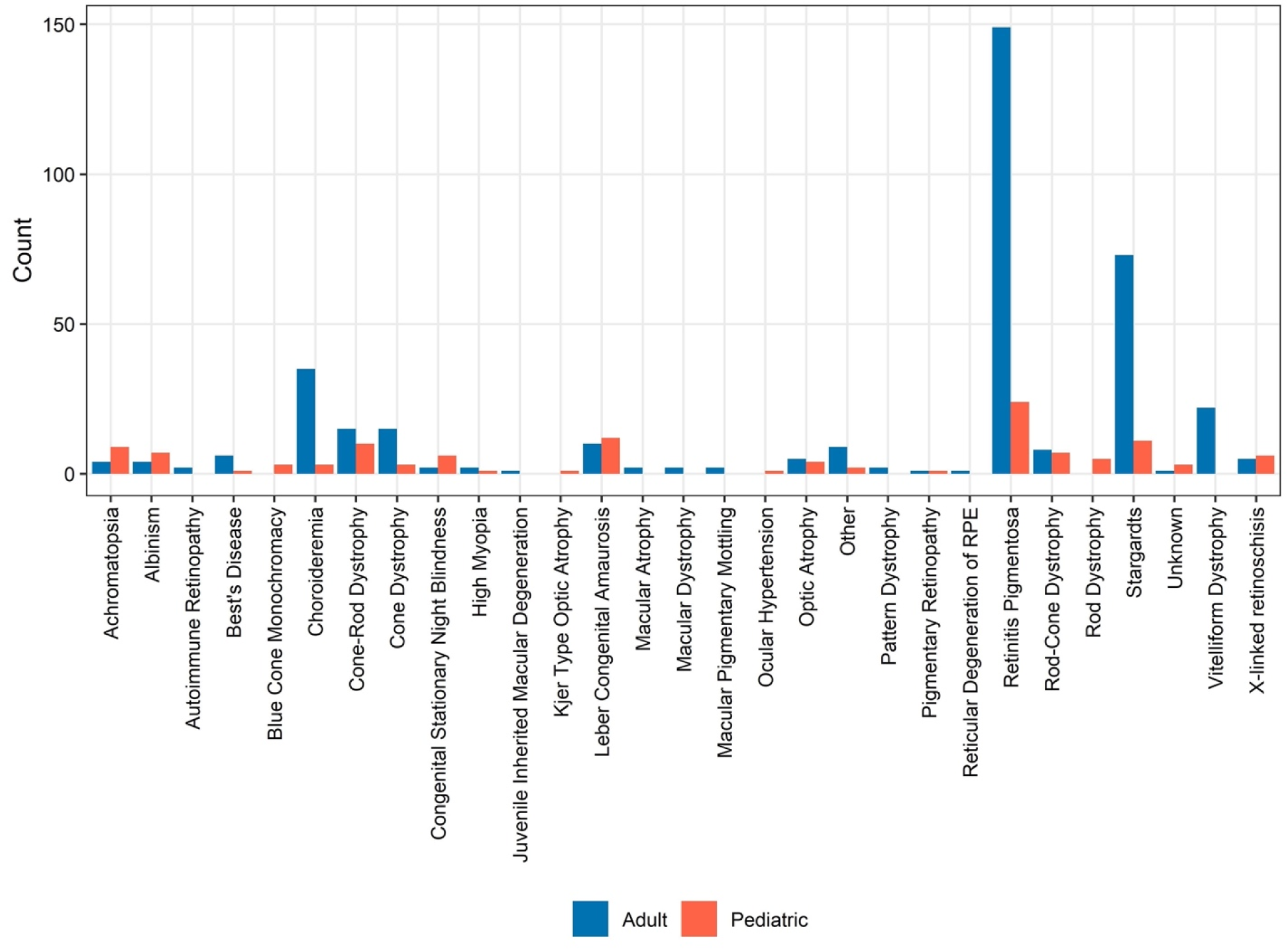
Counts of pediatric and adult patients in relation to ocular diagnosis at time of presentation.

**Table 7.** Post-hoc analysis of proportion of pediatric patients in relation to ocular diagnoses.

| Comparison | *P*_adj |
| --- | --- |
| Choroideremia vs Cone-Rod | 0.004 |
| Choroideremia vs Leber | < 0.001 |
| Choroideremia vs RP | 0.384 |
| Choroideremia vs Stargardts | 0.613 |
| Choroideremia vs Vitelliform | 0.384 |
| Cone-Rod vs Leber | 0.613 |
| Cone-rod vs RP | 0.005 |
| Cone-Rod vs Stargardts | 0.007 |
| Cone-Rod vs Vitelliform | < 0.001 |
| Leber vs RP | < 0.001 |
| Leber vs Stargardts | < 0.001 |
| Leber vs Vitelliform | < 0.001 |
| RP vs Stargardts | 0.710 |
| RP vs Vitelliform | 0.081 |
| Stargardts vs Vitelliform | 0.174 |

## Discussion

This retrospective cohort study of 527 patients with IRDs identified characteristics that were most predictive of a pediatric or adult presentation to a tertiary care center. Previously high myopia has been strongly associated with IRDs^3,4,5,6,7,8^ which our study found as well in line with our hypothesis. Our results indicated that high hyperopia was very strongly associated with IRD presentation in pediatric patients which is less represented in the literature although has been associated^2,4,9^. Similar to previous literature^6^, we found a strong association between astigmatism and IRD. Our finding of a history of developmental delay as a predictor of pediatric IRD makes sense in the context that children who have poor vision are at greater risk for developmental delay as they have limited cues regarding the world around them^12^. Additionally, other studies report developmental delay in some syndromic IRDs^13,14^. Anatomic variants such as polydactyly, brachydactyly and syndactyly have been described in those with IRDs^15^ which correlates with our results of syndactyly as a predictor of IRD in children.

In adults, we found flashes/floaters, reduced central vision, peripheral vision loss, cataracts, diabetes mellitus, obesity, cardiac conditions, and a family history of cataract most predictive of presentation to the IRD clinic. In a large study of patients with RP, 170 patients reported light flashes as a common problem^16^ similar to our findings. The predictors of reduced central and peripheral vision align with previous literature describing the finding of patients with RP becoming legally blind by 40 years of age due to severely reduced visual fields and central visual loss by 60 years of age^17^. Our finding of cataracts predictive of adult presentation aligns well with previous literature. Based on absolute counts, the majority of the adult patients in our study were diagnosed with RP which has been associated with cataract development. In a study by Fishman et al., half of the sample of RP patients were found to have a cataract^5^. We additionally found a family history of cataract to be associated with adult presentation to the IRD clinic which could be explained given the genetic component of IRDs. We found that adults presented to the IRD clinic with medical characteristics including diabetes mellitus, obesity, and a cardiac condition. Previous research has linked syndromic IRDs with diabetes mellitus, obesity, and cardiac problems^18^ which may help to explain our findings. This study further adds to our knowledge of differences in predictors of pediatric versus adult presentation of IRDs.

In contrast to our hypothesis that closer proximity to a tertiary care center would yield a greater proportion of pediatric patients, we found the opposite. A smaller proportion of pediatric patients living closer (<20 miles) to the tertiary care center were found as compared to living farther away. Consequently, pediatric patients presenting with IRDs were more likely to have originated from 20 or more miles away. Having access to a tertiary care center did not lead to more pediatric referrals, and there may be other factors influencing this finding. Perhaps parents are willing to travel farther for their children or perhaps transportation represents a barrier for some patients.

In terms of the type of referring provider, there was no difference between pediatric and adult patients. Based on raw data, the highest number of referrals came from ophthalmologists followed by primary care physicians. This may be because primary care physicians may be an initial point of contact for patients and ophthalmologists are specifically trained in signs to recognize IRDs. As non-eye care professionals encompassed a sizeable portion of referrals, it is important to provide education to clinicians on symptoms that would prompt a patient for referral for an IRD.

The diagnoses of most prevalent IRDs presenting in pediatric versus adult patients in this study reflect current knowledge regarding age of presentation of IRDs^19,20^. For example, our findings indicated a higher proportion of pediatric patients diagnosed with LCA in pediatric patients and a larger number of diagnoses of vitelliform maculopathy at adult onset based on absolute numbers of patients. The larger absolute number of diagnoses of RP correlates to RP encompassing the majority of IRDs with an estimated prevalence of 1 in 4000 individuals^21^. Our study sample reflects prevalence patterns in the adult and pediatric groups.

Limitations of the study include analysis of patients only at a single site instead of multiple sites. Considering the limitations, the study encompassed a large sample size of 527 patients with many data points. This is considerable given that IRDs do not have a high prevalence in the population.

In conclusion, we identified characteristics predictive of pediatric and adult presentation in IRD patients thus addressing current knowledge gaps. It is important to recognize characteristics predictive of IRD presentation to start treatment early and reduce loss of vision. A better understanding of these characteristics may provide for quicker recognition, education of those most likely to encounter IRDs, and allow for earlier treatment. Proximity to a tertiary care center is not the primary factor affecting the age of presentation. Further research is needed to explore barriers for patients from longer distances and outreach strategies for early recognition and referral of IRDs. Furthermore, of interest is exploration of genetic patterns in different regions and communities.

## Data Availability

All data produced in the present study are available upon reasonable request to the authors

## Acknowledgments

Mr. McLaughlin, Mr. Ganansky, Mr. Taher, Ms. Trudrung, Dr. Van De Car, Dr. Le, Dr. Peterson, Dr. Stepien, and Dr. Schmitt have no acknowledgements.

